# Conceptualizing bias in EHR data: A case study in performance disparities by demographic subgroups for a pediatric obesity incidence classifier

**DOI:** 10.1101/2024.02.06.24302390

**Authors:** Elizabeth A. Campbell, Saurav Bose, Aaron J. Masino

## Abstract

Electronic Health Records (EHRs) are increasingly used to develop machine learning models in predictive medicine. There has been limited research on utilizing machine learning methods to predict childhood obesity and related disparities in classifier performance among vulnerable patient subpopulations. In this work, classification models are developed to recognize pediatric obesity using temporal condition patterns obtained from patient EHR data. We trained four machine learning algorithms (Logistic Regression, Random Forest, XGBoost, and Neural Networks) to classify cases and controls as obesity positive or negative, and optimized hyperparameter settings through a bootstrapping methodology. To assess the classifiers for bias, we studied model performance by population subgroups then used permutation analysis to identify the most predictive features for each model and the demographic characteristics of patients with these features. Mean AUC-ROC values were consistent across classifiers, ranging from 0.72-0.80. Some evidence of bias was identified, although this was through the models performing better for minority subgroups (African Americans and patients enrolled in Medicaid). Permutation analysis revealed that patients from vulnerable population subgroups were over-represented among patients with the most predictive diagnostic patterns. We hypothesize that our models performed better on under-represented groups because the features more strongly associated with obesity were more commonly observed among minority patients. These findings highlight the complex ways that bias may arise in machine learning models and can be incorporated into future research to develop a thorough analytical approach to identify and mitigate bias that may arise from features and within EHR datasets when developing more equitable models.

**Author Summary:** Childhood obesity is a pressing health issue. Machine learning methods are useful tools to study and predict the condition. Electronic Health Record (EHR) data may be used in clinical research to develop solutions and improve outcomes for pressing health issues such as pediatric obesity. However, EHR data may contain biases that impact how machine learning models perform for marginalized patient subgroups. In this paper, we present a comprehensive framework of how bias may be present within EHR data and external sources of bias in the model development process. Our pediatric obesity case study describes a detailed exploration of a real-world machine learning model to contextualize how concepts related to EHR data and machine learning model bias occur in an applied setting. We describe how we evaluated our models for bias, and considered how these results are representative of health disparity issues related to pediatric obesity. Our paper adds to the limited body of literature on the use of machine learning methods to study pediatric obesity and investigates the potential pitfalls in using a machine learning approach when studying social significant health issues.

## Introduction

Throughout most disciplines, massive amounts of data are being digitally generated, collected, and stored at a rapidly expanding pace. Additionally, advances in computational methods enable extraction of information from such datasets that produce useful insights and knowledge. (1)

In healthcare, there is increasing use of large and variable data sources that include medical imaging, wearable devices, genome sequencing, and payer records among others; electronic health records (EHRs) are one particularly robust source of healthcare data. This data is available in an exceptionally high volume, spans the healthcare sector’s digital ethos, and is extremely variable in its structure, semantics, and information content. (2,3)

Advanced data mining and analytical methods are necessary to obtain, transform, and analyze EHR data for secondary uses such as clinical and health policy research; machine learning methods are key to addressing challenges in secondary EHR uses. However, although EHR data and the models that may be trained with this data, hold tremendous potential to transform clinical care and research, caution must be exercised in how this data is utilized and interpreted analytically. (4,5) Bias is an inherent property to statistical models and within data collection, and can also be introduced algorithmically or found within the data used to train and test machine learning models.(6)

Issues in using machine learning methods to analyze EHR data often arise when letting data speak for itself. Algorithms may be subject to biases that are present in EHR datasets from several sources including study population characteristics, systemic errors in how EHR data is collected, missing data, misclassification, and sample size. (7) Spurious correlations and other dataset deficiencies such as multicollinear, correlated predictors may lead to algorithms overfitting predictions to biased data and producing unstable estimates. In turn, this affects the models’ performance and generalizability, potentially causing or perpetuating health system disparities. Machine learning models may be subject to new biases not typically seen in more traditional observational studies or statistical methods, such as adjusting away healthcare quality differences between patients or misinterpreting treatment outcomes when making therapy recommendations.(8,9)

### Bias Definitions

Bias that is present in EHR data may result from numerous sources including measurement errors or selection bias in populations that are represented in EHR data versus the communities that they represent. (10) Biased data may reflect existing prejudices or disparities of the contexts from which data are collected. For example, inadequate access to insurance or under-diagnosis of certain conditions may lead to a misrepresentation of a condition’s prevalence among vulnerable populations. In EHR data, these pernicious biases may also manifest from inequities in usage and access to care or in the care that vulnerable subgroups may receive in healthcare settings. Bias may also be introduced algorithmically or in the model design processes, which makes measuring bias when evaluating machine learning models an important area for promoting equity. (11,12) In Figure 1, we conceptualize these EHR data bias sources and how they contribute to developing biased machine learning models in clinical research. In this study, we focus on contextualizing pernicious bias in a particular dataset and how such biases may be characterized.

**Figure 1.**
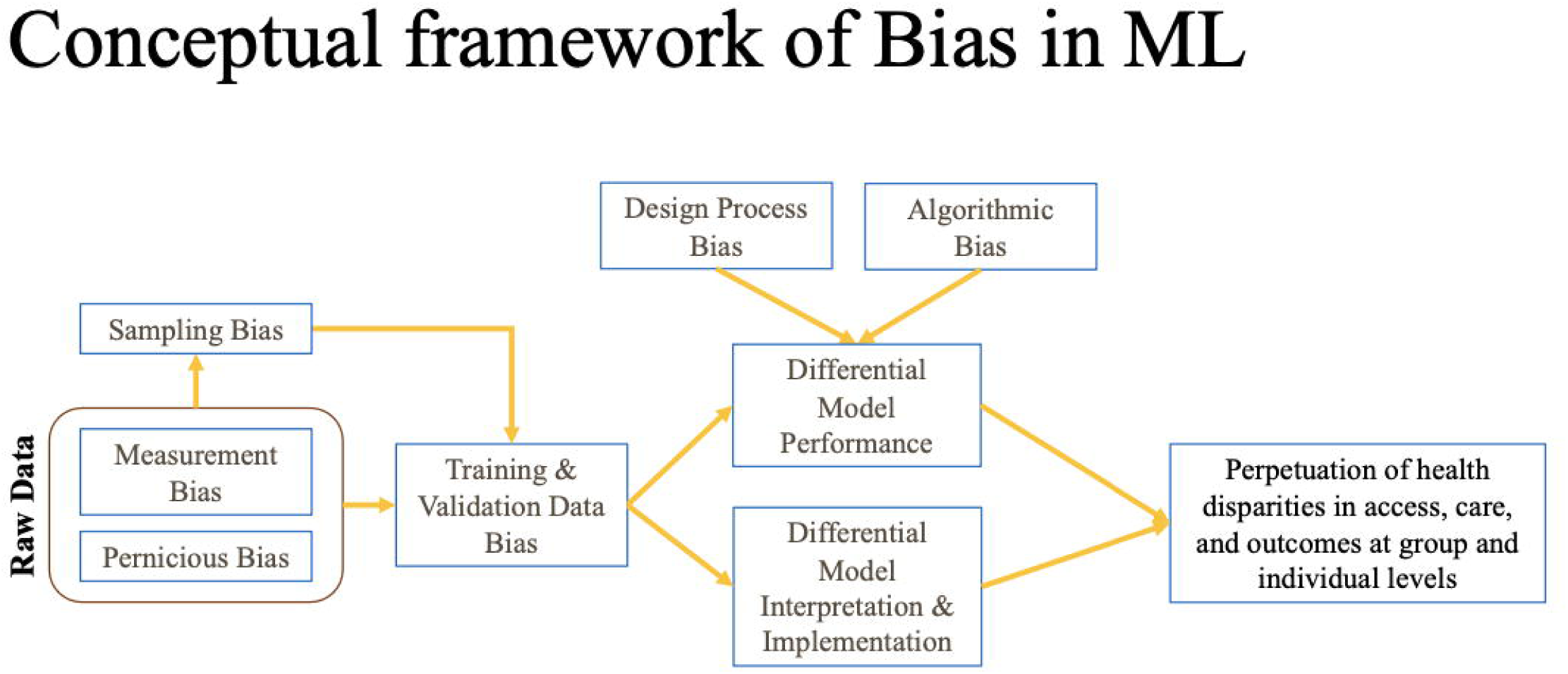
A Framework to Understand Bias in EHR Data and Machine Learning Models. We identify three sources of bias (pernicious, measurement, and sampling biases) that may occur in raw data that translates to biased datasets. Biased data along with bias introduced from design processes and algorithmically may lead to differential machine learning model performance, interpretation, and implementation, which in turn may perpetuate health system disparities.

In this work, we investigate potential biases (which we define here to mean systemic errors or misrepresentations embedded within datasets) that may exist in machine learning models developed from EHR data in the context of a childhood obesity incidence case study. Within the United States, approximately one third of children are overweight (age- and sex-specific body mass index (BMI) greater than or equal to the 85th percentile per Centers for Disease Control and Prevention (CDC) growth charts) or obese (age- and sex-specific BMI greater than or equal to the 95th percentile per CDC growth charts). (13,14) Obesity is linked with numerous comorbidities, including an increased risk of developing asthma, diabetes, hypertension, and psychological conditions during childhood and later in life. (15,16) Pediatric obesity is a socially significant health issue that disproportionately impacts American Indian, African American, and Latino children, compared to non-Hispanic whites. Obesity prevalence is also higher among low-income, rural, or less-educated population subgroups. (13,17)

We developed a classification model to predict childhood obesity incidence using our previously published temporal condition patterns surrounding pediatric obesity that were derived from EHR data and patient demographic data. (18) Our study aims to address the following research questions:

1. Can a machine learning classification model using temporal condition patterns and demographic information from EHR data accurately predict future childhood obesity incidence?
2. How can machine learning models developed using EHR data be analyzed for bias in model performance amongst population subgroups?
3. How can biased model performance be understood in the context of the individual condition that a researcher is working to address?

## Materials and Methods

### Setting

EHR data was obtained from the Pediatric Big Data (PBD) resource at the Children’s Hospital of Philadelphia (CHOP). Patients in this study were from a retrospective cohort of newly obese patients and matched control patients with a healthy BMI identified in a previous study. (19) The PBD resource was created for secondary research use, and contains health-related information, including demographic, encounter, medication, procedure, and measurement (e.g. vital signs, laboratory results) elements for a large, unselected population of children.

Ethics statement: Non-study personnel extracted all data from the EHR and removed protected health information (PHI) identifiers, except for dates, prior to transfer to the study database. Date information was removed from the analysis dataset used in this study. The CHOP Institutional Review Board approved this study and waived the requirement for consent.

### Temporal Condition Pattern Mining Methodology

In a previous study, (18) we applied a sequential pattern mining algorithm to a dataset from a large retrospective cohort of newly obese pediatric patients (n = 49 694) at CHOP from 2009-2017. Patients were identified using the CDC definition of childhood obesity (BMI z-score at or above the 95th percentile for age and sex). (13,14) The BMI z-scores were centrally calculated in this analysis. The same definition of obesity was used across study sites for the entire study period. Patients had at least one obesity measurement during a CHOP primary care visit and at least one visit prior to the first obesity measurement where an obese BMI was not recorded. The analysis aimed to identify common temporal condition patterns derived from visits immediately before (pre-index) and after (post-index) the first visit with an obese BMI (index). We found 163 condition patterns present in at least 1% of the obese patients, of which 80 were more significantly more common than in matched controls. Campbell, et al includes a full study diagram detailing the inclusion criteria implementation for obtaining the study population and derivation of the temporal condition patterns. (18)

### Study Population

To obtain the study population for the machine learning case study presented here, we began with 49,694 pairs of matched cases and controls from the prior study. Patients in our final study population must have had both a BMI measurement in the pre-index and index visit (for control patients the index visit was the date for the visit that they were matched on with their corresponding case patient). For case patients, this meant that they needed to have a non-obese BMI measurement in the pre-index visit, and an obese BMI measurement in the index visit; 15,522 case patients met these criteria. Control patients needed a healthy BMI measurement in both their pre- and index visits; 31,366 control patients met this criterion. Finally, only patients from case-control pairs where both patients met the BMI inclusion criteria were kept in the study population; 4,843 case-control pairs met the criteria and 44,851 did not. A total of 9,686 patients met the BMI criterion for inclusion.

Patients and their corresponding matched case or control were eliminated if they did not have insurance information within 2 years of the matched index visit. For controls, 45 were missing insurance information from within two years or altogether; these 45 controls and their matched cases were eliminated from the study population. Seven cases were missing this information and were eliminated from the study population (along with their matched controls). The final study population contained 4,777 matched pairs, and 9,554 total patients. The study population and data acquisition process are summarized in Figure 2. Table 1 presents the demographic characteristics of the total study population, as well as the case and control populations respectively.

**Figure 2.**
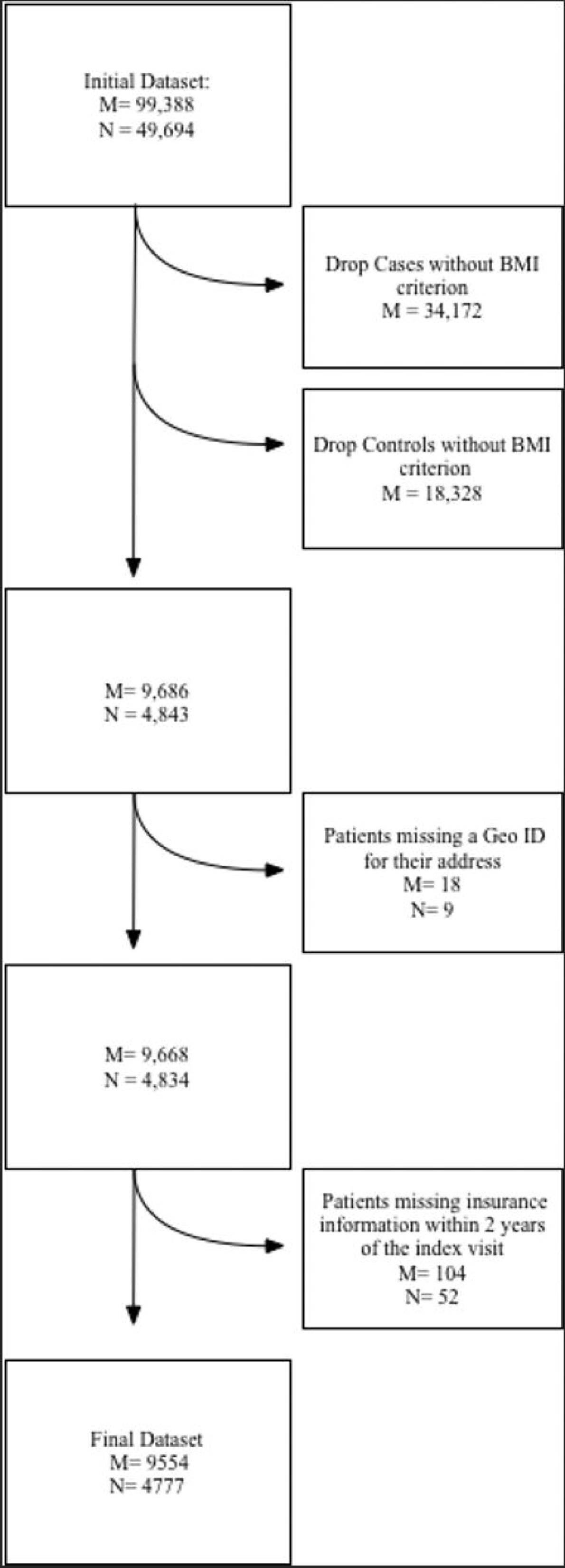
A flow chart illustrating how patients in the final study population were obtained after filtering through the study’s inclusion criteria. M represents the total number of patients, and N represents the total number of matched case-control pairs.

**Table 1.**
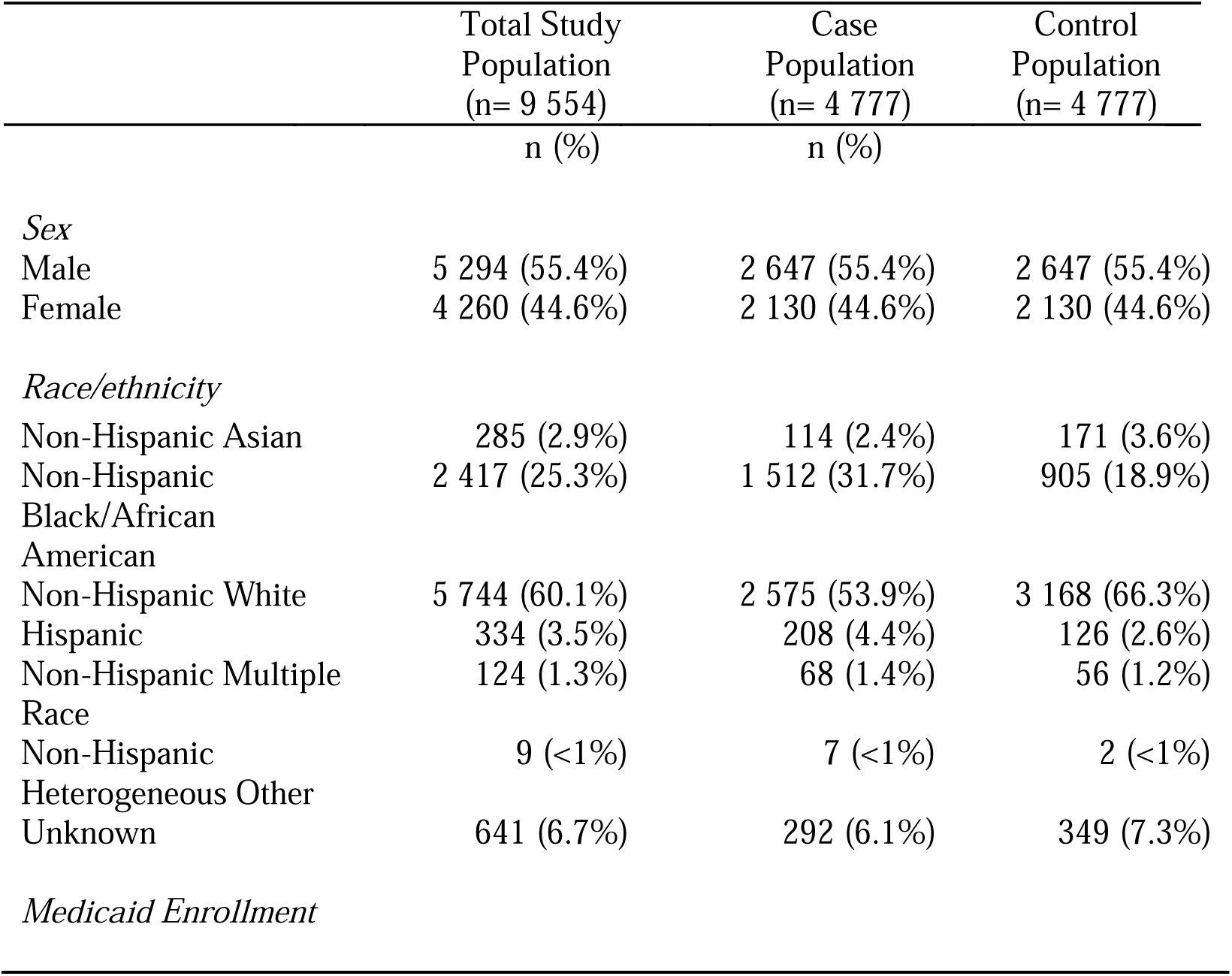
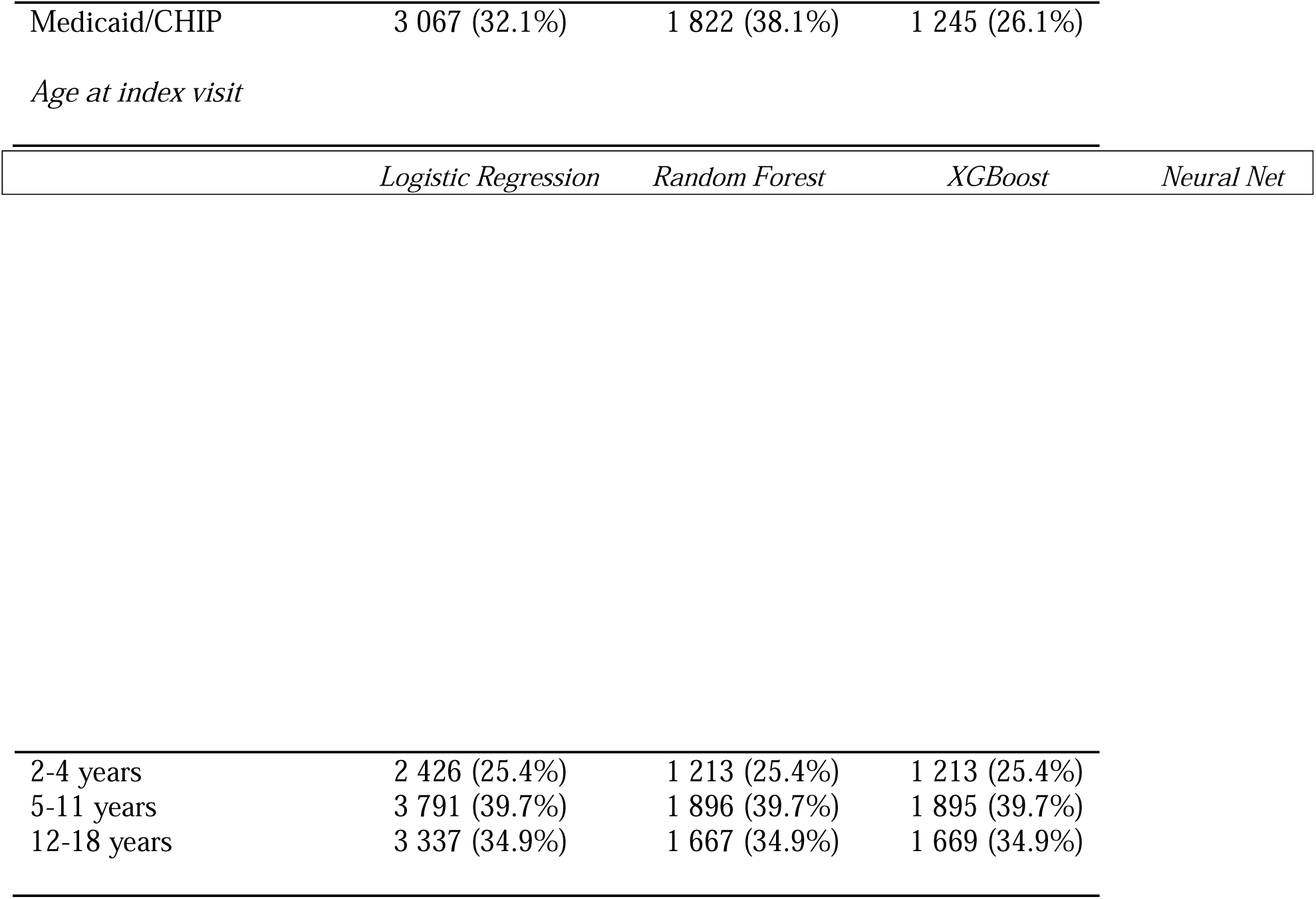
Demographic Characteristics of Obesity Incidence Study Case and Control Populations.

**Table 2.** Mean(SD) AUC-ROC for Study Population and Demographic Subgroups by Classification Algorithm.

|  |  |  |  |  |
| --- | --- | --- | --- | --- |
| *Total Study Population* | 0.78 (0.01) | 0.77 (0.01) | 0.78 (0.01) | 0.76 (0.01) |
| *Sex* |  |  |  |  |
| Male | 0.78 (0.01) | 0.77 (0.01) | 0.78 (0.01) | 0.76 (0.01) |
| Female | 0.78 (0.01) | 0.77 (0.01) | 0.78 (0.01) | 0.77 (0.01) |
| *Race* |  |  |  |  |
| Asian | 0.76 (0.05) | 0.76 (0.05) | 0.77 (0.05) | 0.74 (0.06) |
| Black/African American | 0.79 (0.01) | 0.79 (0.01) | 0.79 (0.02) | 0.78 (0.02) |
| White | 0.75 (0.04) | 0.75 (0.04) | 0.76 (0.01) | 0.74 (0.01) |
| Hispanic | 0.75 (0.04) | 0.75 (0.04) | 0.77 (0.04) | 0.74 (0.04) |
| Multiple Race | 0.73 (0.08) | 0.73 (0.08) | 0.76 (0.08) | 0.75 (0.07) |
| Unknown | 0.73 (0.03) | 0.73 (0.03) | 0.73 (0.04) | 0.72 (0.04) |
| *Medicaid Enrollment* |  |  |  |  |
| Medicaid/CHIP | 0.80 (0.01) | 0.79 (0.01) | 0.80 (0.01) | 0.79 (0.01) |
| Not Enrolled in Medicaid/CHIP | 0.76 (0.01) | 0.75 (0.01) | 0.76 (0.01) | 0.74 (0.01) |
| *Age at index visit* |  |  |  |  |
| 2-4 years | 0.76 (0.02) | 0.75 (0.01) | 0.76 (0.02) | 0.75 (0.01) |
| 5-11 years | 0.80 (0.01) | 0.79 (0.01) | 0.80 (0.01) | 0.79 (0.01) |
| 12-18 years | 0.75 (0.02) | 0.75 (0.02) | 0.76 (0.01) | 0.75 (0.02) |

### Feature Selection, Data Acquisition and Preprocessing

The features selected for the machine learning case study included temporal condition patterns uncovered in Campbell, et al. (18) For obesity incidence prediction, only temporal condition patterns from the pre-index and index visits were considered in this model. Of the original 80 patterns identified in the previous study, 70 temporal condition patterns were selected for inclusion in this analysis. Each temporal condition pattern is considered separately as a feature for this study. Patient EHR data were analyzed for the presence of each temporal condition pattern, and patients were assigned a binary value of 0 (indicating a patient did not have a record of the temporal condition pattern) or a 1 (indicating that a patient did have a record of the temporal condition pattern) for each variable. Diagnoses in the temporal condition patterns are coded using the Expanded Diagnostic Clusters (EDCs) from the Adjusted Clinical Group (ACG) System. (20,21) The temporal diagnoses that comprised the condition patterns used in this study may be found in Table 1 in the Supporting Information section.

Person-level characteristics including race, sex, ethnicity, age at index visit, and insurance were also extracted from EHR data within the PBD database and included in the final dataset. The demographic variables considered were sex assigned at birth, race, Medicaid enrollment (a proxy for socioeconomic status at the time of obesity incidence), (22,23) and age at index visit (coded as a categorical variable, for patients who were 2-5 years, 6-11 years, and 12-18 years). Patients were classified as Hispanic if their self-identified ethnicity was specified as Hispanic or Latino; otherwise, they were categorized by the value of their self-identified race from the EHR. Patients with missing race and ethnicity information were classified as unknown. Patients were classified as being enrolled in Medicaid if they used multiple insurance plans and one of those was Medicaid or Children’s Health Insurance Program (CHIP), Pennsylvania’s state program to provide health insurance to uninsured children and teens who are ineligible or not enrolled in Medicaid. (24) For patients who did not have insurance information recorded for their index visit, all insurance information for their visits within a year of their index visit was obtained from the PBD database and analyzed. If patients had a record of Medicaid/CHIP enrollment within a year of their index visit, they were classified in the Medicaid/CHIP enrollment category.

### Machine Learning Analysis

We trained four machine learning models (Logistic Regression, Random Forest, XGBoost, and Neural Networks) to classify cases and controls as obesity positive or negative, and optimized hyperparameter settings through a bootstrapping methodology. We randomly shuffled the data and split it into training and validation folds in a stratified fashion relative to the 50:50 class balance. We trained each model with all hyperparameter settings on the training fold and evaluated its Area Under the Receiver Operating Curve (AUC-ROC) on the validation fold. We repeated the process 200 times to obtain 200 validation AUC-ROCs for each hyperparameter setting for each model, then selected the hyperparameter combination with the highest mean validation AUC-ROC for a given model class. We implemented all algorithms using the Scikit-learn library in Python 3. (25) We calculated the mean and standard deviation (SD) AUC-ROC values for the total study population and demographic subgroups for each algorithm.

### Permutation Analysis

We performed a permutation feature importance analysis on the data split with median validation AUC-ROC for the model with hyperparameters corresponding to the highest mean validation AUC-ROC. The feature importance is computed by measuring the change in the AUC-ROC on the validation set when the values in the dataset for a given feature are randomly shuffled among samples. Feature importance is reflected by a decrease in AUC-ROC as compared to when the feature is not permuted, with higher importance indicated by a larger decrease.

## Results

### Study Population

Table 1 presents the demographic characteristics of the total study population, as well as the case and control populations respectively.

The study population is majority male (55.4%) and majority White (60.1%). African Americans are the second largest racial /ethnic group (25.3%). Approximately 1/3 of patients (32.1%) were enrolled in Medicaid at the time of their index visit. The case population is majority male (55.4%) and majority White (53.9%) but is comprised of a higher proportion of African Americans (31.7% vs. 25.3%) and Hispanic patients (4.4% vs. 3.5%) patients compared to the entire study population. Additionally, a greater proportion of case patients (38.1%) were enrolled in Medicaid compared to the overall study population (32.1%). The control population has a higher proportion of White patients compared to the case population (66.3% versus 53.9%) and a lower proportion of racial minorities. The control population also had a lower rate of Medicaid enrollment than the case population (26.1% versus 38.1%).

### Machine Learning Results

Mean AUC-ROC values were consistent across algorithms, ranging from 0.72-0.80. On the full study population, Neural Net had a mean AUC-ROC value of 0.76, and mean AUC-ROC values ranged from 0.70-0.79 across demographic subgroups. On the full study population, Random Forest had a mean AUC-ROC value of 0.77, and mean AUC-ROC values ranged from 0.73-0.79 across demographic subgroups. On the full study population, Logistic Regression had a mean AUC-ROC value of 0.77, and mean AUC-ROC values ranged from 0.73-0.80 across demographic subgroups. On the full study population, XGBoost had a mean AUC-ROC value of 0.78, and mean AUC-ROC values ranged from 0.73-0.80 across demographic subgroups. XGBoost and Logistic regression tended to perform the best on the full study population and when evaluated by demographic subgroups. Some evidence of bias was identified, although surprisingly this was through the models performing better for minority subgroups. The highest mean AUC-ROC value (0.80) was observed among African American patients, patients enrolled in Medicaid, and patients ages 5-11 years.

### Permutation Analysis Findings

A permutation analysis was undertaken to investigate why models tended to perform slightly better for under-represented groups in the study population. We hypothesized that the features that are most predictive of obesity may be more common among marginalized subpopulations. Thus, we undertook a permutation feature analysis to identify which features were most important in classifying patients as obese or not obese for each algorithm.

Table 3 presents the top ten most predictive sequences for each classification algorithm. Four temporal condition patterns were among the top ten most predictive features across all four algorithms: 1-ALL04 (a diagnosis of asthma in the pre-index visit), 2-MUS01 (a diagnosis of Musculoskeletal signs and symptoms in the index visit), 2-ALL03 (a diagnosis of allergic rhinitis in the index visit0, and 2-SKN04 (a diagnosis of acne in the index visit). A diagnosis of asthma in the pre-index visit (1-ALL04) was the most predictive feature for the XGBoost, Neural Network, and Random Forest algorithms, and was the third most predictive for Logistic Regression. To better understand the disparate machine learning model performance and permutation analysis findings, the prevalence of these four temporal condition patterns were assessed among demographic subgroups in the study population, Table 4.

**Table 3.** Most Predictive Variables by Classifier (Value (2 sigma)) The top ten most predictive sequences for each classification algorithm. The gray highlighted cells represent sequences that were most predictive across all four classifiers. The orange highlighted cells indicate race variables that were most predictive.

| Variable Predictive Rank | Classifier<br>XGBoost | Random Forest | Neural Net | Logistic Regression |
| --- | --- | --- | --- | --- |
| 1 | 1-ALL04<br>2.04% (.37%) | 1-ALL04<br>2.10% (.60%) | 1-ALL04<br>1.13 % (.57%) | race_3<br>1.52 % (.55%) |
| 2 | 2-MUS01<br>1.36% (.32%) | 2-ALL03<br>1.41% (.65%) | 2-MUS01<br>0.93% (.30%) | 2-MUS01<br>1.26% (.28%) |
| 3 | 2-ALL03<br>1.15% (.25%) | 2-NUR19<br>1.31% (.62%) | 1-ALL03, 1-ALL04<br>0.85% (.35%) | 1-ALL04<br>1.11% (.41%) |
| 4 | 2-SKN04<br>0.83% (.29%) | race_2<br>1.05% (.41%) | 1-SKN02<br>0.79% (.34%) | 2-MUS04<br>0.99% (.20%) |
| 5 | 1-SKN02<br>0.79% (.33%) | 1-SKN02<br>0.82% (.40%) | 2-ALL03<br>0.79% (.40%) | 2-NUR19<br>0.97% (.41%) |
| 6 | 2-END05<br>0.75% (.18%) | 2-MUS01<br>0.82% (.32%) | race_3<br>0.70% (.62%) | 1-ALL03, 1-ALL04<br>0.87% (.20%) |
| 7 | race_2<br>0.72% (.26%) | 2-RES01<br>0.77% (.37%) | 2-SKN04<br>0.61% (.22%) | 2-RES01<br>0.69% (.41%) |
| 8 | 2-ALL04<br>0.72% (.41%) | 1-GAS03<br>0.70% (.54%) | 2-ALL04<br>0.60% (.86%) | 2-ALL03<br>0.67% (.50%) |
| 9 | 2-MUS17<br>0.70% (.11%) | 2-ALL04<br>0.65% (.47%) | 1-GAS03<br>0.60% (.22%) | 2-SKN04<br>0.66% (.39%) |
| 10 | 2-MUS04<br>0.68% (.31%) | 2-SKN04<br>0.59% (.27%) | 2-RES01<br>0.54% (.45%) | medicaid<br>0.55% (.29%) |

**Table 4.**
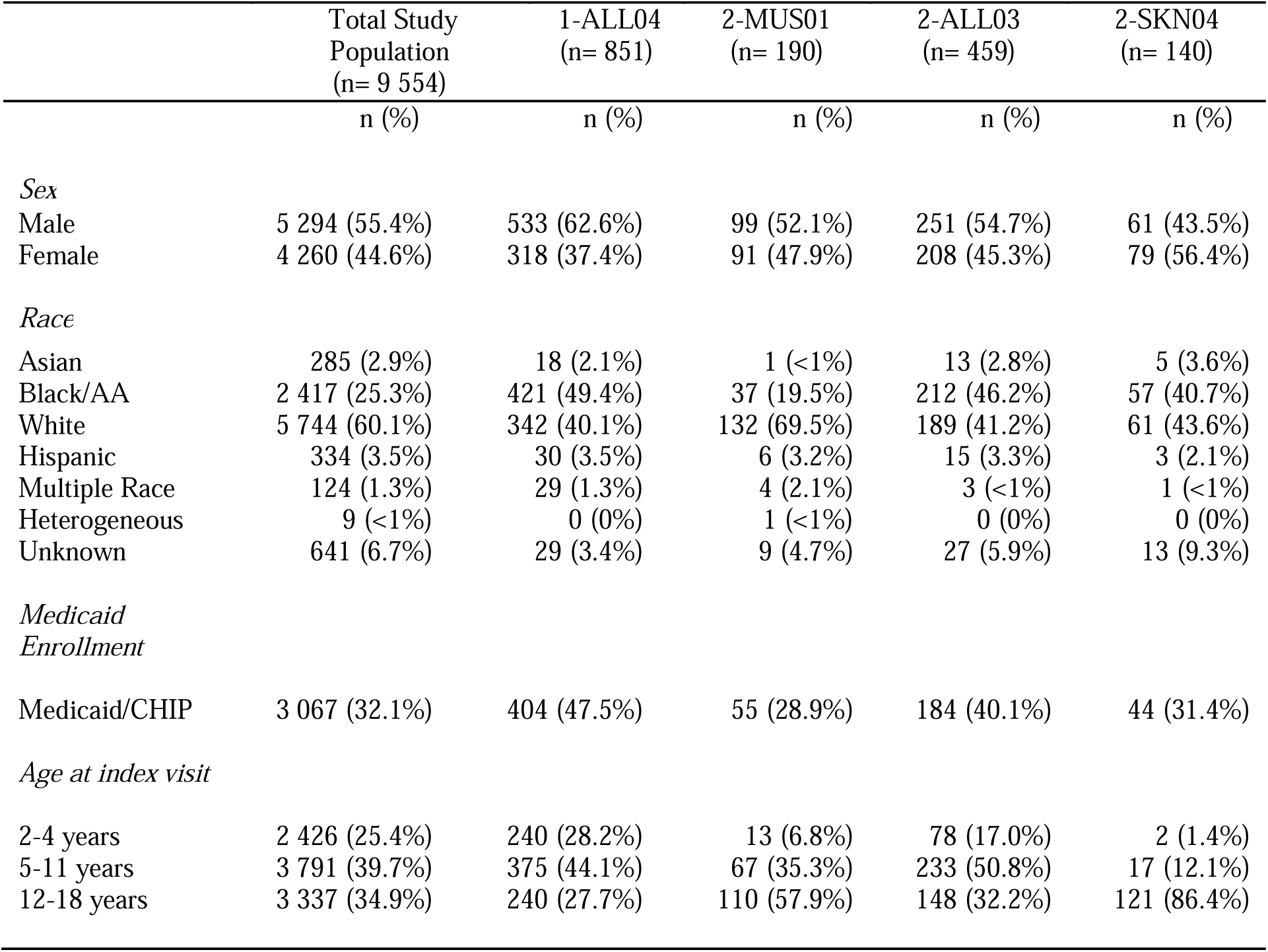
Demographic Characteristics of Patient Subgroups with most predictive sequences.

Two features that were most common among patients were also a most predictive sequence for all four classifiers: 1-ALL04 (asthma in the pre-index visit), of which 851 patients had a record in their EHR data, and 2-ALL03 (allergic rhinitis in the index visit) of which 459 patients had a record (compared to 190 patients with the 2-MUS01 sequence and 140 patients with the 2-SKN04 diagnosis). African American patients and patients enrolled in Medicaid are over-represented among patients who have these diagnoses. Almost half of patients (49.4%) with a 1-ALL04 diagnosis were African American, even though African American patients make up only 25.3% of the study population, and 47.5% of patients with this diagnosis were enrolled in Medicaid compared to only 32.1% of the total study population. Similarly, 46.2% of patients with the 2-ALL03 diagnosis were African American and 40.1% were enrolled in Medicaid.

## Discussion

In this study, four supervised machine learning algorithms were trained to identify pediatric patients as obese or not obese using demographic variables and temporal condition patterns previously found to be associated with obesity incidence. Model performance was evaluated for the total population and by demographic subgroups using mean AUC-ROC values. Mean AUC-ROC values were consistent across algorithms, ranging from 0.72-0.80. XGBoost and Logistic regression tended to perform the best on the full study population and when evaluated by demographic subgroups. Algorithms tended to perform relatively consistently compared to one another and when each classifier’s performance was analyzed by demographic subgroups. This result is consistent with existing work demonstrating that complex machine learning models often perform no better than logistic regression when using EHR input data. (26) Our conjecture is that this is likely true when inputs consist of highly informative structured data such as that found in the EHR and similarly, the temporal patterns used in our work.

A permutation analysis was conducted on the classifiers developed in our case study. The ten variables that were most predictive for each of the four classifiers developed in the obesity incidence prediction research were identified and their average impact for each model’s AUC-ROC was computed. Two variables, 1-ALL04 (asthma in the pre-index visit) and 2-ALL03 (allergic rhinitis in the index visit), were among the most predictive variables for all four classifiers and were the most prevalent condition patterns among the study population. African American patients and patients enrolled in Medicaid were over-represented among patients who had this temporal condition pattern. Our findings align with prior research on the association between asthma and pediatric obesity which provide insight into why these variables were so informative. Pediatric obesity and asthma are strongly associated, and early-life asthma contributes to the onset of pediatric obesity.(27) Although the relationship between allergic rhinitis and pediatric obesity is unclear, (28,29) allergic rhinitis has been shown to be comorbid with pediatric asthma.(30) Low-income, urban, and racial minority children are disproportionately impacted by both pediatric obesity and asthma, (17,31,32) which explains their over-representation amongst patients with the most predictive features and the slight classifier performance bias in their favor.

Some evidence of model bias relative to population subtypes was detected, although unexpectedly the bias manifested as the classifiers tending to perform better on vulnerable subgroups including African American patients and patients enrolled in Medicaid (a proxy for lower socioeconomic status) than the entire population. These findings illustrate that there are many complex ways that bias may emerge within EHR data, and that the context in which data is collected and population impacted by a condition must be carefully considered when assessing EHR data for bias. Prior work has shown that bias in machine learning typically results in lower model performance for minorities due to an under-representation in data. (33,34) However, possible biases need to be examined carefully. We hypothesize that the features most predictive of obesity are more represented among patients in under-represented subgroups in this study, which lends to the classification algorithms performing generally equitably if not slightly better for African American patients and patients enrolled in Medicaid (a bias that is in favor of vulnerable subpopulations). While these are simply associations and causality cannot be inferred, our results support the idea that causes of bias in datasets and the models trained from them are much more nuanced than initially thought.

### Limitations

While this study serves to illustrate challenges and nuances associated with bias in machine learning models developed using EHR, it does have limitations. First, the temporal condition patterns mined from EHR data that were utilized as features for the machine learning models only show associations. Findings are descriptive and the discovered temporal patterns and comorbidities should be viewed in this light. No causality can be attributed to the associations uncovered in this study. Similarly, the comparisons made in classifier performance differences between population subgroups and the prevalence of temporal condition pattern prevalence are also descriptive. No tests of statistical significance were performed, as this was a descriptive case study that represents a first step in further research into bias in machine learning models developed from EHR data. Finally, we acknowledge that when considering the outcome of obesity in the machine learning prediction problem, minority patients comprised a greater proportion of the case population compared to controls. This may have contributed to the model performance bias in favor of vulnerable subgroups.

### Conclusion

Our paper presents a comprehensive framework of how bias may be present within EHR data and external sources of bias in the model development process, which in turn impacts machine learning model development and clinical applications. Our pediatric obesity case study describes a detailed exploration of a real-world machine learning model to contextualize how concepts related to EHR data and machine learning model bias occur in an applied setting. We describe how we evaluated our models for bias, and considered how these results are representative of health disparity issues related to pediatric obesity. Finally, our paper presents a novel application of data-driven temporal condition patterns that surround pediatric obesity incidence into a predictive machine learning model. This adds to the limited body of literature on the use of machine learning methods to study pediatric obesity and investigates the potential pitfalls in using a machine learning approach when studying social significant health issues.

Bias is a complex and multi-faceted issue that is present in society and translates into data collected in applied settings. We expect that our study may be used to define the types of bias that researchers working with EHR data to develop machine learning models may look for, and to understand that bias may manifest in machine learning models in unexpected ways. Our approach to evaluating a machine learning model for bias and contextualizing our model evaluation alongside clinical and psychosocial knowledge surrounding pediatric obesity provides a useful blueprint for researchers developing and evaluating machine learning models with EHR data in the obesity space and beyond. Finally, our findings support more equitable model development, and may be used to guide researchers and clinicians in the precision medicine space to consider the types of bias that may be present in machine learning models and how to implement these models in clinical settings in a way that helps to address and not advance existing systemic disparities.

## Contributors

EC, SB, and AM contributed to the study’s conception and design. EC conducted the study’s literature review and drafted the paper. EC and SB contributed to data acquisition, dataset development, and data analysis. EC, SB, and AM contributed to interpretation of findings. AM obtained funding to support this study and supervised study implementation. EC, SB, and AM revised the paper. All authors approved the final version of the manuscript. All authors had full access to the data in the study and were involved in data interpretation and writing of the report. All authors had final responsibility for the decision to submit for publication.

## Data sharing

Data cannot be shared publicly because of HIPAA requirements. Please contact with questions regarding data availability.

## Acknowledgements

This work was supported by a grant from the Commonwealth Universal Research Enhancement (C.U.R.E.) program funded by the Pennsylvania Department of Health—2015 Formula award— SAP #4100072543. This work was also supported by funding from The Children’s Hospital of Philadelphia (CHOP)-Drexel Research Fellowship Program: Informatics and Analytics Collaborative Research. We would like to thank the investigators of the Pediatric Big Health Data initiative for their contributions. These individuals include: Christopher B. Forrest, MD, PhD; L. Charles Bailey, MD, PhD; Shweta P. Chavan, MSEE; Rahul A. Darwar, MPH; Daniel Forsyth; Chén C. Kenyon, MD, MSHP; Ritu Khare, PhD; Mitchell G. Maltenfort, PhD; Xueqin Pang, PhD; Hanieh Razzaghi, MPH; Justine Shults, PhD; Levon H. Utidjian, MD, MBI from the Children’s Hospital of Philadelphia; Ana Diez Roux, MD, PhD, MPH; Amy H. Auchincloss, PhD, MPH; Kimberly Daniels, MS; Anneclaire J. De Roos, PhD, MPH; J. Felipe Garcia-Espana, MS, PhD; Irene Headen, PhD, MS; Félice Lê-Scherban, PhD, MPH; Steven Melly, MS, MA; Yvonne L. Michael, ScD, SM; Kari Moore, MS; Abigail E. Mudd, MPH; Leah Schinasi, PhD, MSPH from Drexel University and, Yong Chen, PhD; John H. Holmes, PhD; Rebecca A. Hubbard, PhD; A. Russell Localio, JD, MPH, PhD from the University of Pennsylvania.

## Supporting Information

**S1 Table.**
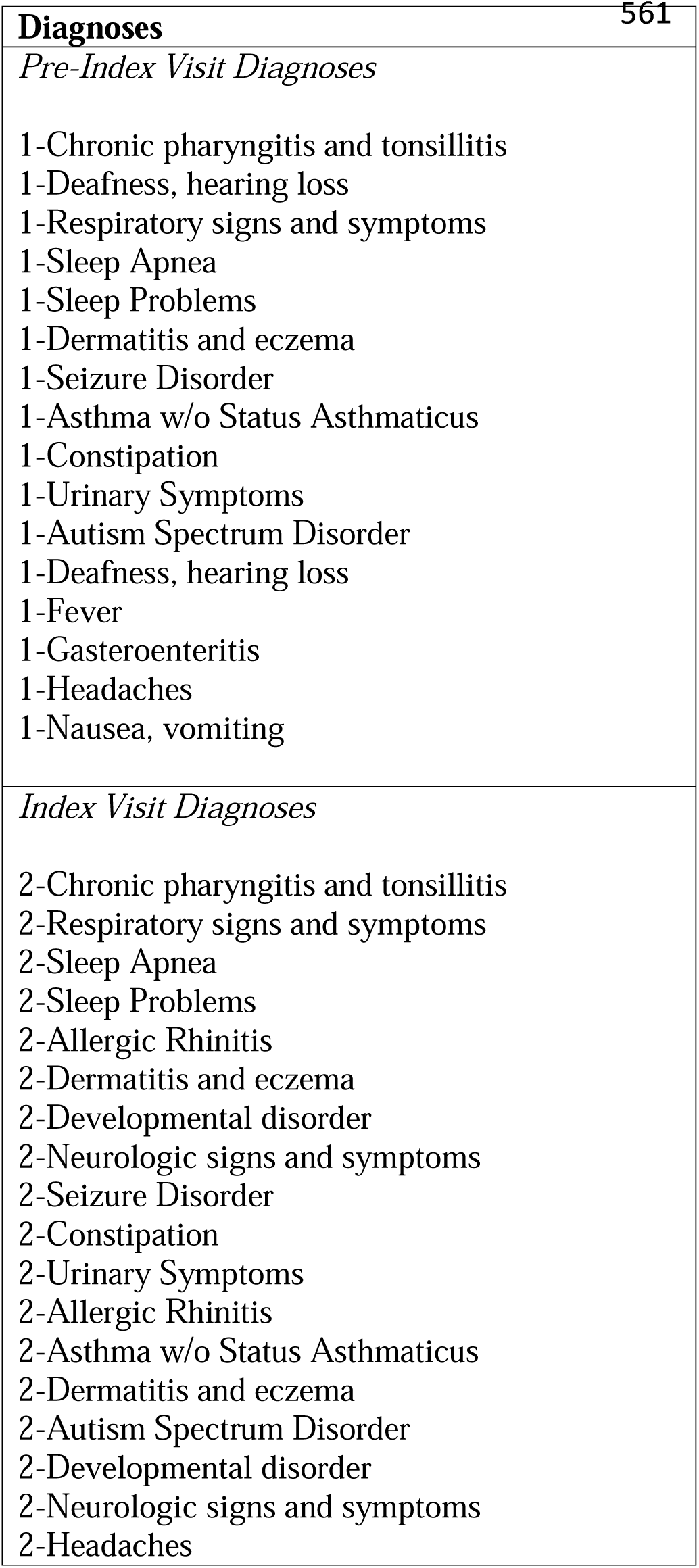
Temporal Diagnoses Included as Machine Learning Model Features.

## References

1. Murdoch TB, Detsky AS. The Inevitable Application of Big Data to Health Care. JAMA. 2013 Apr 3;309(13):1351–2.

2. J. Andreu-Perez, C. C. Y. Poon, R. D. Merrifield, S. T. C. Wong, G. -Z. Yang. Big Data for Health. IEEE Journal of Biomedical and Health Informatics. 2015 Jul;19(4):1193–208.

3. Ferrao JC, Oliveira MD, Janela F, Martins HM. Preprocessing structured clinical data for predictive modeling and decision support. Applied clinical informatics. 2016;7(04):1135–53.

4. Char DS, Shah NH, Magnus D. Implementing machine learning in health care—addressing ethical challenges. The New England journal of medicine. 2018;378(11):981.

5. Carter SM, Rogers W, Win KT, Frazer H, Richards B, Houssami N. The ethical, legal and social implications of using artificial intelligence systems in breast cancer care. The Breast. 2020;49:25–32.

6. Shah DS, Schwartz HA, Hovy D. Predictive Biases in Natural Language Processing Models: A Conceptual Framework and Overview. In 2020. p. 5248–64.

7. Yadav P, Steinbach M, Kumar V, Simon G. Mining electronic health records (EHRs) A survey. ACM Computing Surveys (CSUR). 2018;50(6):1–40.

8. Gianfrancesco MA, Tamang S, Yazdany J, Schmajuk G. Potential biases in machine learning algorithms using electronic health record data. JAMA internal medicine. 2018;178(11):1544–7.

9. Auerbach A, Fihn SD. Discovery, Learning, and Experimentation With Artificial Intelligence–Based Tools at the Point of Care—Perils and Opportunity. JAMA Network Open. 2021;4(3):e211474–e211474.

10. Cutillo CM, Sharma KR, Foschini L, Kundu S, Mackintosh M, Mandl KD. Machine intelligence in healthcare—perspectives on trustworthiness, explainability, usability, and transparency. NPJ digital medicine. 2020;3(1):1–5.

11. McCradden MD, Joshi S, Anderson JA, Mazwi M, Goldenberg A, Zlotnik Shaul R. Patient safety and quality improvement: Ethical principles for a regulatory approach to bias in healthcare machine learning. Journal of the American Medical Informatics Association. 2020;27(12):2024–7.

12. Chen IY, Szolovits P, Ghassemi M. Can AI help reduce disparities in general medical and mental health care? AMA journal of ethics. 2019;21(2):167–79.

13. Skinner AC, Ravanbakht SN, Skelton JA, Perrin EM, Armstrong SC. Prevalence of obesity and severe obesity in US children, 1999–2016. Pediatrics. 2018;141(3).

14. Kuczmarski RJ. 2000 CDC Growth Charts for the United States: methods and development. Department of Health and Human Services, Centers for Disease Control and …; 2002.

15. Karnik S, Kanekar A. Childhood obesity: a global public health crisis. Int J Prev Med. 2012;3(1):1–7.

16. Pulgarón ER. Childhood obesity: a review of increased risk for physical and psychological comorbidities. Clinical therapeutics. 2013;35(1):A18–32.

17. Ogden CL, Carroll MD, Fakhouri TH, Hales CM, Fryar CD, Li X, et al. Prevalence of obesity among youths by household income and education level of head of household— United States 2011–2014. Morbidity and mortality weekly report. 2018;67(6):186.

18. Campbell EA, Qian T, Miller JM, Bass EJ, Masino AJ. Identification of temporal condition patterns associated with pediatric obesity incidence using sequence mining and big data. International Journal of Obesity. 2020;44(8):1753–65.

19. Campbell EA, Bass EJ, Masino AJ. Temporal condition pattern mining in large, sparse electronic health record data: A case study in characterizing pediatric asthma. Journal of the American Medical Informatics Association. 2020;27(4):558–66.

20. Bailey LC, Milov DE, Kelleher K, Kahn MG, Del Beccaro M, Yu F, et al. Multi-institutional sharing of electronic health record data to assess childhood obesity. PloS one. 2013;8(6):e66192.

21. Weiner J, Abrams C. The Johns Hopkins ACG System Technical Reference Guide, Version 10.0. John Hopkins Bloomberg School of Public Health. 2011;

22. Arpey NC, Gaglioti AH, Rosenbaum ME. How socioeconomic status affects patient perceptions of health care: a qualitative study. Journal of primary care & community health. 2017;8(3):169–75.

23. Schechter MS, Shelton BJ, Margolis PA, FitzSimmons SC. The association of socioeconomic status with outcomes in cystic fibrosis patients in the United States. American journal of respiratory and critical care medicine. 2001;163(6):1331–7.

24. About CHIP: Commonwealth of Pennsylvania [Internet]. 2019. Available from: https://www.chipcoverspakids.com/AboutCHIP/Pages/default.aspx

25. Pedregosa F, Varoquaux G, Gramfort A, Michel V, Thirion B, Grisel O, et al. Scikit-learn: Machine learning in Python. the Journal of machine Learning research. 2011;12:2825–30.

26. Christodoulou E, Ma J, Collins GS, Steyerberg EW, Verbakel JY, Van Calster B. A systematic review shows no performance benefit of machine learning over logistic regression for clinical prediction models. Journal of clinical epidemiology. 2019;110:12–22.

27. Azizpour Y, Delpisheh A, Montazeri Z, Sayehmiri K, Darabi B. Effect of childhood BMI on asthma: a systematic review and meta-analysis of case-control studies. BMC pediatrics. 2018;18(1):1–13.

28. Sidell D, Shapiro NL, Bhattacharyya N. Obesity and the risk of chronic rhinosinusitis, allergic rhinitis, and acute otitis media in school_Jage children. The Laryngoscope. 2013;123(10):2360–3.

29. Weinmayr G, Forastiere F, Büchele G, Jaensch A, Strachan DP, Nagel G, et al. Overweight/obesity and respiratory and allergic disease in children: international study of asthma and allergies in childhood (ISAAC) phase two. PloS one. 2014;9(12):e113996.

30. Stern J, Chen M, Fagnano M, Halterman JS. Allergic rhinitis co-morbidity on asthma outcomes in city school children. Journal of Asthma. 2022;1–7.

31. Thakur N, Oh SS, Nguyen EA, Martin M, Roth LA, Galanter J, et al. Socioeconomic status and childhood asthma in urban minority youths. The GALA II and SAGE II studies. American journal of respiratory and critical care medicine. 2013;188(10):1202–9.

32. Persky VW, Slezak J, Contreras A, Becker L, Hernandez E, Ramakrishnan V, et al. Relationships of race and socioeconomic status with prevalence, severity, and symptoms of asthma in Chicago school children. Annals of Allergy, Asthma & Immunology. 1998;81(3):266–71.

33. Obermeyer Z, Powers B, Vogeli C, Mullainathan S. Dissecting racial bias in an algorithm used to manage the health of populations. Science. 2019;366(6464):447–53.

34. Zhang H, Lu AX, Abdalla M, McDermott M, Ghassemi M. Hurtful words: quantifying biases in clinical contextual word embeddings. In 2020. p. 110–20.

